# Task-based fMRI in early Multiple Sclerosis: what is the best head motion correction approach?

**DOI:** 10.1101/2022.01.31.22270167

**Authors:** Júlia F. Soares, Rodolfo Abreu, Ana Cláudia Lima, Lívia Sousa, Sónia Batista, Miguel Castelo-Branco, João Valente Duarte

## Abstract

Functional MRI (fMRI) is one of the most common brain imaging modalities used for understanding brain organization and connectivity abnormalities associated with multiple sclerosis (MS). The fMRI signal is highly perturbed by head motion, which degrades data quality and influences all image-derived metrics. Numerous correction approaches have been proposed over the years to overcome the problems induced by head motion, however, despite a few efforts, there are still current and persistent controversies regarding the best correction strategy. The lack of a systematic comparison between different correction approaches motivates the search for optimal correction models, particularly in studies with clinical populations prone to characterize by higher motion. Moreover, motion correction strategies gain more relevance in task-based designs, which are less explored compared to resting-state and may have a crucial role in describing the functioning of the brain and highlighting specific connectivity changes.

We acquired fMRI data from a group of patients with early MS and matched healthy controls (HC) during performance of a visual task, characterized motion in both groups, and compared the most used motion correction methods. We compared task-activation metrics obtained from models without motion correction, models containing 6 or 24 motion parameters (MPs) as nuisance regressors, models containing 6 or 24 MPs and motion outliers detected with FD or DVARS as nuisance regressors (scrubbing) and models with 6 or 24 MPs where motion outliers were corrected through volume interpolation. To our knowledge, volume interpolation is a frequently used approach but was never compared with other existent methods.

Our results showed that there were no differences in motion between groups, suggesting that recently diagnosed MS patients do not present problematic motion. In general, models with 6 MPs present higher Z-scores than models with 24 MPs, suggesting the 6 MPs as the best trade-off between motion correction and preservation of valuable information. However, correction approaches differ between groups, regarding the combination of MPs with correction of motion outliers. Models with 6 MPs and outliers’ volume interpolation or scrubbing with FD presented higher Z-scores in the MS group, while models with 6 MPs and scrubbing with DVARS or volume interpolation were the best combinations for the HC group. Differences between groups in motion correction strategies draw attention to the intrinsic impact of MS on fMRI analyses, which should be carefully addressed.

This work paves the way towards finding an optimal motion correction strategy, which is required to improve the accuracy of fMRI analyses, crucially in clinical studies in MS and other patient populations.

## 1. Introduction

Resting-state functional MRI (rs-fMRI) has evolved to become one of the most common brain imaging modalities and has been critical for understanding fundamental properties of brain organization and connectivity abnormalities associated with diverse clinical conditions (Binnewijzend et al., 2012; Roosendaal et al., 2010; Schoonheim et al., 2010; van Duinkerken et al., 2012). Particularly, multiple sclerosis (MS) is a disconnection disease that is due to structural damage but also functional connectivity alterations, which has been extensively investigated with fMRI during rest (Eijlers et al., 2019; Meijer et al., 2020; Sbardella et al., 2015; Shu et al., 2016). However, task-designs target brain regions and networks that show distinct properties than in resting-state (Di et al., 2013; Schoonheim et al., 2015). Thus, task-fMRI may have a crucial role in describing the functioning of the brain, in highlighting specific connectivity changes, and thus in understanding this disease better. However, the blood oxygen-level-dependent (BOLD) signal measured with fMRI is highly susceptible to various sources of noise.

Head motion is the main source of noise in the BOLD signal. Motion artifacts degrade data quality and influence all image-derived metrics such as task activation and connectivity estimates (Liu, 2016; Zeng et al., 2014). On the one hand, rs-fMRI studies have demonstrated that head motion can introduce systematic bias to connectivity estimates by creating spurious but spatially structured patterns in functional connectivity (Maknojia et al., 2019; Parkes et al., 2018; Power et al., 2014). On the other hand, in task-based fMRI studies, head motion is more problematic when it correlates with the experimental tasks leading to false brain activations. If not properly accounted for, head motion will bias the statistical results, reducing the sensitivity and specificity for detecting task-specific BOLD responses (Caballero-Gaudes and Reynolds, 2017; Power et al., 2014; Seto et al., 2001). To obtain a “clean” signal with neuronal and biological validity is then important to mitigate the effects of head motion. This is crucial in studies with developmental or clinical populations, especially those that tend to move more, where diagnosis and monitoring need to be the most accurate as possible (Griffanti et al., 2016; Saccà et al., 2021). It has been reported that early diagnosed MS patients and patients with higher disability levels tend to move, while at rest, to a greater extent in the MRI scanner than control subjects (Boonstra et al., 2017; Saccà et al., 2019, 2018). Also, a task-based fMRI study has found a linear increase in motion as task difficulty increased that was larger among MS patients with lower cognitive ability (Wylie et al., 2014). Furthermore, activation in the sensory-motor cortex during performance of a complex bilateral finger tapping task was also found to be greater in control subjects compared to relatively healthy MS patients, as a consequence of head motion in MS (Lowe et al., 2006). However, the effects of head motion in task-fMRI studies of MS, especially in early stages where head motion can be less evident but still present, and considering other task designs, are not systematically explored. Namely, there is a lack of studies investigating and comparing the efficacy of different methods to mitigate the impact of motion artifacts on fMRI measures, particularly in the context of MS. Apart from ensuring the participant remains as still as possible within the MRI scanner, there are a plethora of methods described in the literature to correct head motion effects in data processing. However, there is no consensus over the most appropriate approach (Zaitsev et al., 2015). In fact, a systematic evaluation of motion correction approaches, in resting-state, has shown an heterogeneous efficacy of different methods and suggest that different strategies may be appropriate depending on the context (Ciric et al., 2017). Due to the growing interest in using task activation studies and since these are more prone to motion artifacts compared to resting state studies, we focused this work on systematically comparing some of the most used strategies to correct head motion in a task-based fMRI study with patients with multiple sclerosis and healthy controls.

To apply the proper correction method, it is important to acknowledge the type of head motion we are dealing with. There are two main types: gradual head shifts, and sudden movements of the head known as motion outliers. The most common approach to compensate for head shifts, which is common in all fMRI preprocessing pipelines, is to realign all fMRI volumes to a reference volume, usually the first or the middle volume from the fMRI time series (Power et al., 2015). The position of the head in space is estimated at each volume relatively to the reference volume using rigid body transformations where the head position is described at each timepoint by six motion parameters (MPs): translational displacements along X, Y, and Z axes; and rotational displacements of pitch, yaw, and roll. Then, these 6 MPs can be included as nuisance regressors in a General Linear Model (GLM) analysis of the fMRI data to account for the variance of the BOLD signal explained by the head shifts. However, because residual BOLD variance associated with head shifts can still be present, additional MP-derived regressors have been suggested although being less commonly used, namely the temporal derivatives of the MPs (Power et al., 2012) and the quadratic terms, resulting in a total set of 12MPs and 24MPs, respectively (Satterthwaite et al., 2013; Turner et al., 1996). Additionally, to gradual head shifts, motion outliers are more problematic and generate the most critical BOLD signal changes (Power et al., 2012). These can be identified as spikes in the data time courses and cause large variations in image intensity. Such spikes are not accurately estimated using rigid body transformations, and thus the realignment step or the regression of the MPs fails to account for them. As a solution, several metrics have been proposed for describing subject motion and the detection of motion outliers, the most common being the Root Mean Squared head position change (RMS movement), the Framewise Displacement (FD), and the Derivative or root mean square VARiance over voxelS (DVARS), with the latter being a particular form of the RMS. (Power et al., 2012) compared the FD and DVARS metrics in terms of movement characterization and found that these provide very similar results, however it was unclear whether one index captures data quality better than the other. In any case, when these summary statistics are above a certain threshold for a particular volume (e.g., values of 0.5 for FD and 0.5% ΔBOLD for DVARS), this volume is considered essentially unusable. Nonetheless, motion outliers can still be corrected through different ways, with the most common being censoring and scrubbing. Censoring is simply removing the outlier volumes most affected by the movement from the data, which might result in biased samples (Parkes et al., 2018). Scrubbing follows a model-driven strategy, whereby the volumes affected by extreme motion are identified and additional scan nulling regressors (with 1s at the volumes where motion spikes are detected and 0s elsewhere) are regressed out from the fMRI (Siegel et al., 2014). These can be either regressed out via GLM where the regressors are added directly in the GLM and accounted as covariates or nuisance regressors, or via multiple regression where the output residuals constitute the signal free of noise. Alternatively, volumes associated with motion outliers can be interpolated based on non-corrupted volumes (Caballero-Gaudes and Reynolds, 2017; Mazaika et al., 2009; Mckechanie et al., 2019; Rudas et al., 2020).

Despite all the worthy efforts, there is still no consensus regarding the optimal number of MP-related regressors to consider for tackling head shifts, nor the most appropriate additional approach to mitigate motion outliers (Zaitsev et al., 2015). In (Mascali et al., 2021), denoising pipelines including realignment/tissue-based regression with 24MPs, principal component analysis (PCA) or independent component analysis (ICA) methods (aCompCor and ICA-AROMA, respectively), global signal regression, and censoring of motion-contaminated volumes were compared for task-based functional connectivity. In (Ciric et al., 2017) the same denoising pipelines plus spike regression (de-spiking) and scrubbing have been compared in a resting state framework, suggesting that different strategies may be appropriate depending on the context. The same methods were evaluated with data from clinical populations in (Parkes et al., 2018). However, the volume interpolation method was not addressed in these studies and comparisons of the same approach but with different motion detection metrics (e.g., FD vs DVARS) were not reported. Furthermore, the comparison of correction strategies in the context of MS is lacking. Activation and connectivity measures during task performance may have a crucial role in describing the functioning of the brain, highlighting specific connectivity changes, and thus understanding better this disease. Thus, it is crucial to investigate the interaction effect of the disease with motion correction strategies, to provide robust measures that might help to understand the pathophysiology of the disease and also serve as a tool for disease assessment of progression, ideally in a real-world clinical scenario.

In this study we aim to characterize head motion and compare the most used correction strategies in clinical context using fMRI data collected from early diagnosed MS patients and healthy control subjects, during the performance of one visual passive task and one (more demanding) visual perceptual decision-making task. We started by computing head motion metrics for the two groups to study if there are relevant differences between early MS patients and controls. Next, we compared the most used strategies to correct the effects of head motion and tested if the group has influence on the choice of the correction method. The strategies we compared are models with 6 and 24 MPs, mainly designed to deal with head gradual movements, and models with 6 or 24 MPs plus methods for tackling motion outliers to investigate if these can provide a better correction than models with only 6 and 24MPs. We compared scrubbing methods with two different motion outliers’ detection metrics, FD and DVARS, and volume interpolation. The best approach was determined based on the quality of the data analyses, given by activation and variance-explained metrics.

## 2. Materials and Methods

### 2.1 Participants

All participants gave written informed consent to participate in the study after a full verbal and written explanation of the study. The study was approved by the ethics committees of the Faculty of Medicine of the University of Coimbra (reference CE-047/2018) and of the CHUC (reference CHUC-048-19), and the study was carried in accordance with the Code of Ethics of the World Medical Association (Declaration of Helsinki) for experiments involving humans. Patients were recruited and clinically assessed at the Neurology Department of the Coimbra Hospital and University Centre (CHUC) and met the criteria for MS diagnosis according to McDonald Criteria (Thompson et al., 2018). This study included 11 patients recently diagnosed with MS and 8 healthy control (HC) subjects. Patients also underwent neuropsychological evaluation with the Brief International Cognitive Assessment for MS (BICAMS) (Langdon et al., 2012). Demographic data are presented in Table 1.

**Table 1:** Demographic data of the participants. “F” stands for female, “R” stands for right. EDSS is the Expanded Disability Status Scale.

| Group | Age | Gender | Handedness | EDSS |
| --- | --- | --- | --- | --- |
| MS | 32.18 ± 8.05 | 6F | 11R | 2.05 ± 0.52 |
| HC | 30.75 ± 8.61 | 4F | 8R | - |

### 2.2 fMRI data acquisition

Imaging was performed at the Portuguese Brain Imaging Network facilities (Coimbra, Portugal) on a 3T Siemens MAGNETOM Prisma Fit MRI scanner (Siemens, Erlangen, Germany) using a 64-channel RF receive coil. fMRI data was acquired using a 2D simultaneous multi-slice (SMS) gradient-echo echoplanar imaging (GE-EPI) sequence (6× SMS and 2×in-plane GRAPPA accelerations), with the following parameters: TR/TE = 1000/37 ms, voxel size = 2.0×2.0×2.0 mm^3^, 72 axial slices (whole-brain coverage), FOV = 200×200 mm^2^, FA = 68°, and phase encoding in the anterior-posterior direction. A short EPI acquisition (10 volumes) with reversed phase encoding direction (posterior-anterior) was also performed prior to each fMRI run, for image geometric distortion correction. A 3D anatomical T1-weighted MP2RAGE (TR = 5000 ms, TE = 3.11 ms; 192 interleaved slices with isotropic voxel size of 1 mm^3^) was also collected for subsequent image registration.

### 2.3 Experimental Protocol

The experimental protocol consisted of three functional runs: one run of a passive visual task, which was a functional localizer of the human middle temporal area (hMT+/V5, a low-level visual area well-known to respond to simple motion patterns), and two runs of a decision-making visual task of biological motion (BM) perception.

The localizer run consisted of 10 blocks of 18 seconds, with each block comprising three periods: the first was a fixation period marked by a red cross positioned at the center of the screen for 6 seconds. During the second period, a pattern of stationary dots was shown for 6 seconds, followed by the third (and final) period during which the dots were moving towards and away from a central fixation cross at a constant speed (5 deg/sec) for 6 seconds.

Biological motion stimuli were built based on human motion capture data collected at 60 Hz, comprising 12 point-lights placed at the main joints of a male walker. Each BM perception run consisted of 12 blocks of 40 seconds: 4 or 5 blocks (depending on the starting block) of the point-light walker facing rightwards or leftwards (global biological motion), 4 or 5 blocks showing only the point-light located at the right ankle and moving rightwards of leftwards (local biological motion), and 3 blocks of point lights randomly positioned across the y axis, while maintaining their true trajectory across the x axis (scrambled motion). A total of 9 global, 9 local and 6 random blocks were presented during the two BM perception runs. After each stimulus presentation, the participants reported the direction of motion of the dots (left or right) by pressing one of two buttons.

### 2.4 fMRI data preprocessing

fMRI data were preprocessed using custom scripts in MATLAB®, using the SPM12 software with CAT12 and PhysIO toolboxes (Kasper et al., 2017), and FMRIB Software Library (FSL). The preprocessing pipeline included: 1) slice timing correction; 2) realignment of all fMRI volumes relative to the first volume; 3) correction of geometric distortions caused by magnetic field inhomogeneity, with FSL tool TOPUP (Andersson et al., 2003) using the reversed-phase encoding acquisition; 4) bias field correction; 5) image registration (functional to structural); 5) segmentation of the T1 structural image (with CAT12 toolbox) to extract WM and ventricular CSF masks; 6) estimation of nuisance regressors (with PhysIO toolbox) such as cardiac and respiratory signals, WM and ventricular CSF average BOLD fluctuations and head motion (6 and 24 MPs and motion spikes); 7) Regression of noise fluctuations. Then the “clean images” from the regression were brain masked and the preprocessing was completed with spatial smoothing with a 3 mm full-width-at-half-maximum (FWHM) isotropic Gaussian kernel and high-pass temporal filtering with a cut-off period of 24s and 80s for the localizer and BM tasks respectively.

### 2.5 Motion quantification

We characterized and compared head motion in both groups. For this characterization we individually computed the typical framewise displacement (FD), *meanFD*, the FD without considering the time series’ volumes affected by motion outliers, *meanFD’*, the FD considering only the time points where motion spikes were detected, *meanFD’’*, the number of spikes, and the amount of variance of the average BOLD signal explained by motion, computed through the 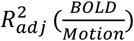 formula.

FD is a scalar quantity to express instantaneous head motion and it is computed through the time series of the 6 MPs obtained during the motion correction step (Power et al., 2012). The FD is expressed by:

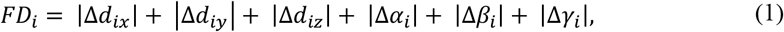

where Δ*d*_*ix*_ = *d*_*(i − 1)x*_ − *d*_*ix*_, and similarly for the other motion parameters, *d*_*ix*_, *d*_*iy*_, *d*_*iz*_, *α*_*ix*_, *β*_*ix*_, *γ*_*ix*_

The FD was obtained with PhysIO toolbox. We computed the FD without considering the motion spikes and the FD considering only the spikes to understand how much the spikes would contribute to degradation of the BOLD signal due to intense movements. The number of spikes was given by the number of points detected by FD with motion above 0.5 mm.

DVARS is a measure computed from the data itself and does not depend on the MPs. It represents how much the intensity of a volume changes in comparison to the previous one (Power et al., 2012). The DVARS metric is given by:

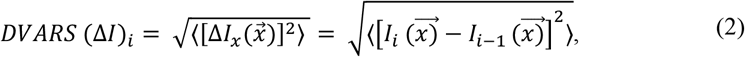

Where 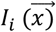 is the image intensity at locus (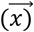) on frame *i* and angle brackets denote the spatial average over the whole brain. The DVARS was computed with FSL tool *fsl_motion_outliers*, and motion outliers were identified by thresholding the DVARS at the 75th percentile plus 1.5 times the inter-quartile range.

We also computed the 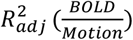 measure as an additional metric to quantify motion between groups, which was estimated by the coefficient of determination adjusted for the degrees of freedom, defined according to (Montgomery et al., 2012):

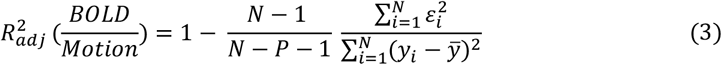

where 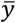 is the average BOLD signal, *N* is the number of volumes and *P* the number of motion regressors; ε denotes the residual of the model under analysis, which is described by *ε* = *y*-*βX*, where *X* is the matrix containing the MPs, and β the associated weights estimated using a GLM framework. For each method (combination of MPs with scrubbing/interpolation) we computed the percentage of variation of the BOLD signal without motion correction explained by the motion regressors. The higher the value of 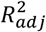. the more variance of the BOLD signal is explained by motion, so the better is the method in capturing and correcting for head motion effects on the data.

Here, we tested 6 and 24 MPs because they represent the two extreme cases complexity-wise (Maknojia et al., 2019). The 6 MPs were obtained during realignment and the 24 MPs which correspond to squares of the 6 MPs and temporal derivatives were obtained with PhysIO toolbox. Then we compared the different correction methods between groups based on quality metrics (described below). The goal is to identify which strategy, among the combination of 6 MPs or 24MPs with scrubbing with FD, scrubbing with DVARS or volume interpolation is better to mitigate the effects of motion. Models with only 6 and 24 MPs were mainly designed to understand which set of MPs is better for dealing with gradual movements. To correct the impact of gradual head motion, 6 MPs and 24 MPs are regressed out from the BOLD signal in the regression step of the preprocessing pipeline. The scrubbing method was implemented by identifying the head spikes, through FD and DVARS with the thresholds mentioned above, with 1’s and 0’s elsewhere in the design matrix. Then these regressors are also regressed out from the BOLD signal in the regression step of the preprocessing pipeline. Volume interpolation was implemented with *ArtRepair* toolbox as the final step of the preprocessing where the affected volumes were firstly identified by FD with a threshold of 0.5mm and then interpolated based on non-corrupted volumes. Finally, a signal free of motion-related noise is ready to be integrated in a General Linear Model (GLM) framework to obtain the statistical maps where the quality metrics will be computed to compare the different correction approaches.

### 2.6 Statistical Analysis

The GLM framework was used to map the regions involved in our perceptual task. It is basically a linear regression represented by:

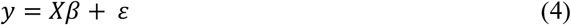

with y the time series from one voxel, *X* the design matrix, *β* the model parameters, *ε*, the normally distributed error (or residuals) with zero mean (Pernet, 2014). Onsets and durations of each experimental condition were included in the model of the BOLD signal as regressors of interest representative of our tasks. For the localizer task we ended up with two regressors representing periods showing static points and moving points whereas for the BM tasks three regressors representing periods showing global biological motion, local biological motion, and scrambled motion were added to the model. These regressors were built based on unit boxcar functions with ones during the respective periods, and zeros elsewhere and convolved with a canonical, double gamma hemodynamic response function (HRF). The HRF-convolved regressors were then included in a GLM that was subsequently fitted to the fMRI data. After the fitting, the β weights are estimated, which represent the relevance of each regressor in explaining the variance of the data. Here, we set out to study brain regions that are activated when visual motion is present. Thus, the areas associated with these conditions were localized according to the contrasts [motion - static] and [global BM motion + local BM motion + scrambled motion - baseline] for the localizer and BM runs, respectively. We used family wise error (FWE) correction for multiple comparisons based on Random Field Theory (RFT), and we only considered activations as significant those with a threshold of *p* < 0.05, with a cluster-level threshold of *p* < 0.05. One GLM was estimated for each participant and for each run, thus each participant ended up with 9 statistical maps per run: i) map with no motion correction. The only preprocessing step related to motion was realignment of the volumes to the first volume of the temporal series. These maps act as control to see how much movement was corrected with the different correction methods; ii) map with 6 MPs; iii) map with 24 MPs; iv) map with 6 MPs and scrubbing with FD; v) map with 6 MPs and scrubbing with DVARS; vi) map with 6 MPs and volume interpolation; vii) map with 24 MPs and scrubbing with FD; viii) map with 24 MPs and scrubbing with DVARS; ix) map with 24 MPs and scrubbing with volume interpolation. From the resulting activation maps, the quality metrics were extracted.

#### 2.6.1 Quality Metrics

The maximum (Z-max) and mean (Z-mean) Z-score values were extracted from each statistical map in each subject. The Z values indicate the sensitivity of the model in detecting brain regions that are associated with our tasks. The higher the values of Z, the higher is the goodness of fit of the GLM explaining data variation, thus higher is the accuracy of the motion correction method. We decided to average the Z-score values of the three runs (one run of localizer task and two runs of BM task) because the Z-score values were very similar across the runs and the regions that activate in each one are identical, as expected because participants performed visual motion tasks in both.

#### 2.6.2 Comparisons

To statistically compare the amount of head motion between groups, a t-test was applied to measures of FD, number of spikes, and 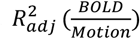. To evaluate the performance of the correction methods tested here, models of analysis of variance with repeated measures for the quality metrics, Z-max and Z-mean, were used. To compare the strategies mostly used to correct the gradual head shifts, a two-way mixed MANOVA (one between-subjects and one within-subjects factor) was performed. Similarly, to compare the strategies for correcting motion outliers’ effects and to study if scrubbing or volume interpolation methods are worth adding to the models with only 6 or 24MPs for correction of gradual shifts, a three-way mixed MANOVA (one between-subjects and two within-subjects factors) was performed. The between-subjects factor in the two comparisons is *Group*, which has two nominal unrelated or independent categories: Multiple Sclerosis (MS) and control (HC) participants. For the first comparison, the within-subjects’ factor is the *MPs* (number of motion parameters), with two levels (6 MPs and 24 MPs) and for the second comparison, the within-subjects’ factors are the *MPs* and motion outliers’ *Correction Method*, with three levels (INTERP, FD and DVARS).

## 3. Results

### 3.1 Motion characterization

Motion characterization, evidencing FD measurements, 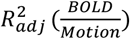 and the number of spikes for both groups, is represented in Figure 1. T-tests for these metrics revealed that no statistically significant differences were found between MS patients and controls (significance threshold, corrected for multiple comparisons, of *p*-value = 0.004):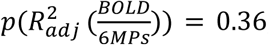; 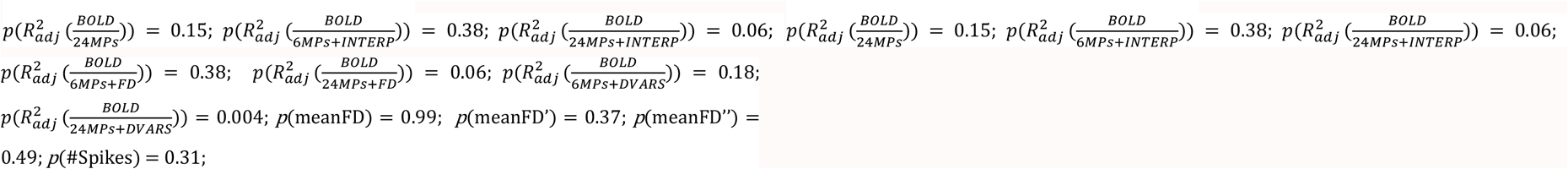

**Figure 1:**
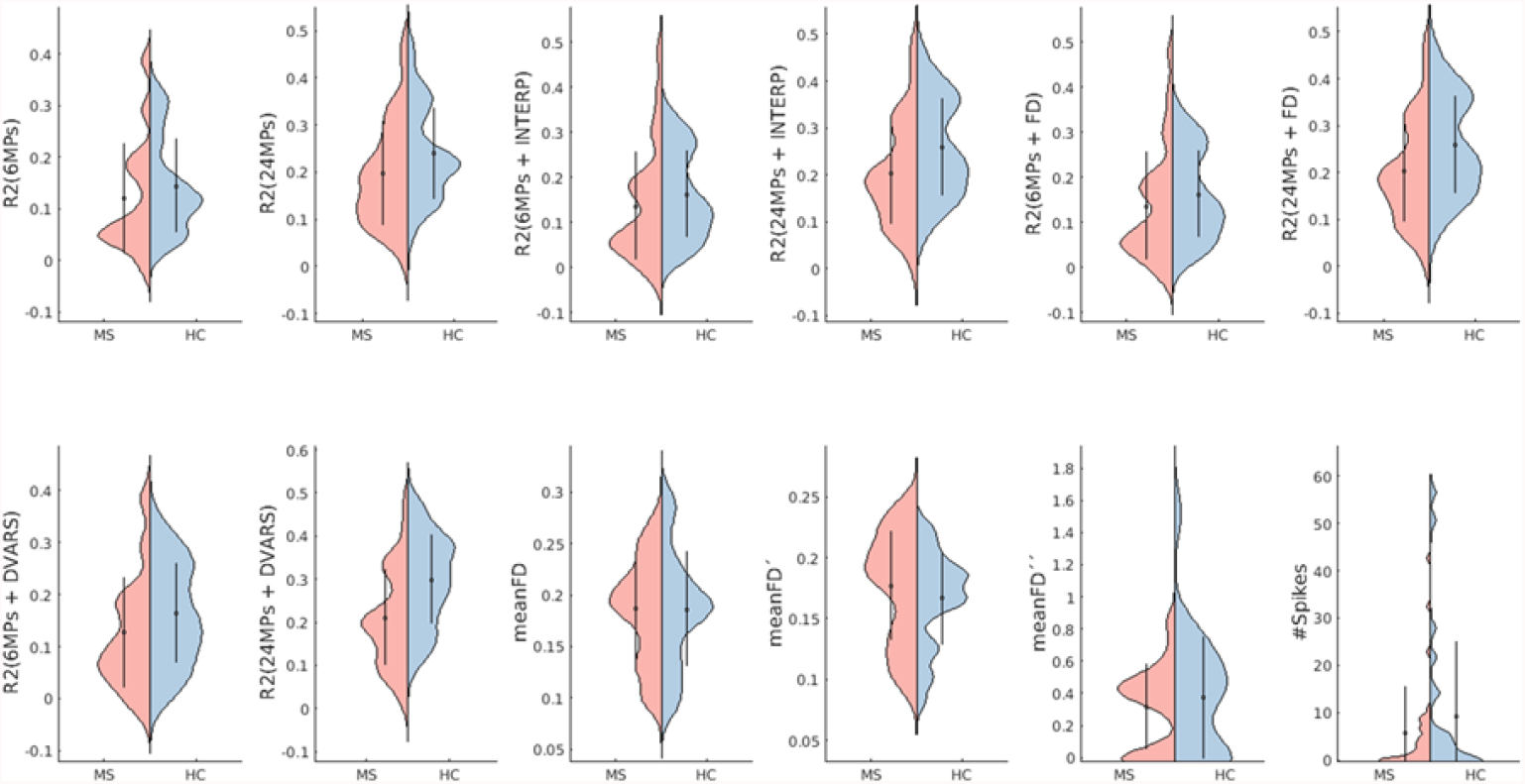
Motion quantification. Violin plots of motion metrics for both groups. R^2^ describes the amount of BOLD signal variation (without motion correction) explained by each set of regressors describing motion. meanFD is the mean framewise displacement, meanFD’ is the mean FD without considering motion outliers, meanFD’’ is the mean FD considering only the motion outliers and #Spikes is the number of motion spikes. Red and blue represent the groups of MS patients and HC, respectively. The dots and vertical lines in each group represent the mean ± standard deviation. Both distributions are quite similar, evidencing no differences in motion metrics between groups, as supported by t-tests.

### 3.2 Correction Methods Comparison

The quality metrics of the models, group mean Z-max and Z-mean, for models with only MPs regressors for correction of gradual shifts are shown in Table 2. In Table 3 we present the metrics for the models with the combination of MPs for correction of gradual shifts and motion outliers’ correction methods.

**Table 2:** Metrics to assess the quality of the models using motion correction of gradual head shifts with 6 MPs and 24 MPs. Values are presented as mean ± standard deviation in each group of participants.

| Correction | Group | Z-max | Z-mean |
| --- | --- | --- | --- |
| 6MPs | MS | 8.55 ± 0.60 | 6.72 ± 0.50 |
| | HC | 7.96 ± 0.89 | 6.30 ± 0.63 |
| 24MPs | MS | 8.26 ± 0.78 | 6.53 ± 0.56 |
| | HC | 7.56 ± 1.07 | 6.10 ± 0.62 |

**Table 3:** Metrics to assess the quality of the models using a combination of 6 MPs or 24 MPs with each method to correct the motion outliers’ effects. Values are presented as mean ± standard deviation in each group of participants. “INTERP” stands for volume interpolation models, “FD” are the models with scrubbing using FD as the outliers’ detection metric, “DVARS” represents the models with scrubbing using DVARS as the outliers’ detection metric.

| Correction | MPs | Group | Metrics |  |
| --- | --- | --- | --- | --- |
|  |  |  | Zmax | Zmean |
| INTERP | 6MPs | MS | 8.54 ± 0.62 | 6.73 ± 0.51 |
| | | HC | 7.99 ± 0.87 | 6.31 ± 0.63 |
| | 24MPs | MS | 8.26 ± 0.78 | 6.54 ± 0.57 |
| | | HC | 7.59 ± 1.05 | 6.12 ± 0.61 |
| FD | 6MPs | MS | 8.53 ± 0.60 | 6.72 ± 0.50 |
| | | HC | 7.85 ± 1.02 | 6.27 ± 0.65 |
| | 24MPs | MS | 8.25 ± 0.77 | 6.52 ± 0.51 |
| | | HC | 7.53 ± 1.10 | 6.07 ± 0.65 |
| DVARs | 6MPs | MS | 8.49 ± 0.61 | 6.69 ± 0.51 |
| | | HC | 8.28 ± 0.75 | 6.55 ± 0.56 |
| | 24MPs | MS | 8.20 ± 0.75 | 6.48 ± 0.56 |
| | | HC | 7.85 ± 0.96 | 6.26 ± 0.57 |

The two-way interaction of the two-way mixed MANOVA was non-significant, *p* = 0.168. Subsequently, the between-subjects’ effect of *Group* was significant (*p* = 0.005 for Z-max and *p* = 0.007 for Z-mean) and the main effect of *MPs* was also significant (*p* < 0.001), with pairwise comparisons showing higher Z-scores for maps with 6 MPs. This means that using 6 MPs is better than using 24 MPs regardless of the group. Figure 2 shows mean activation maps of models containing 6 and 24 MPs for each group.

**Figure 2:**
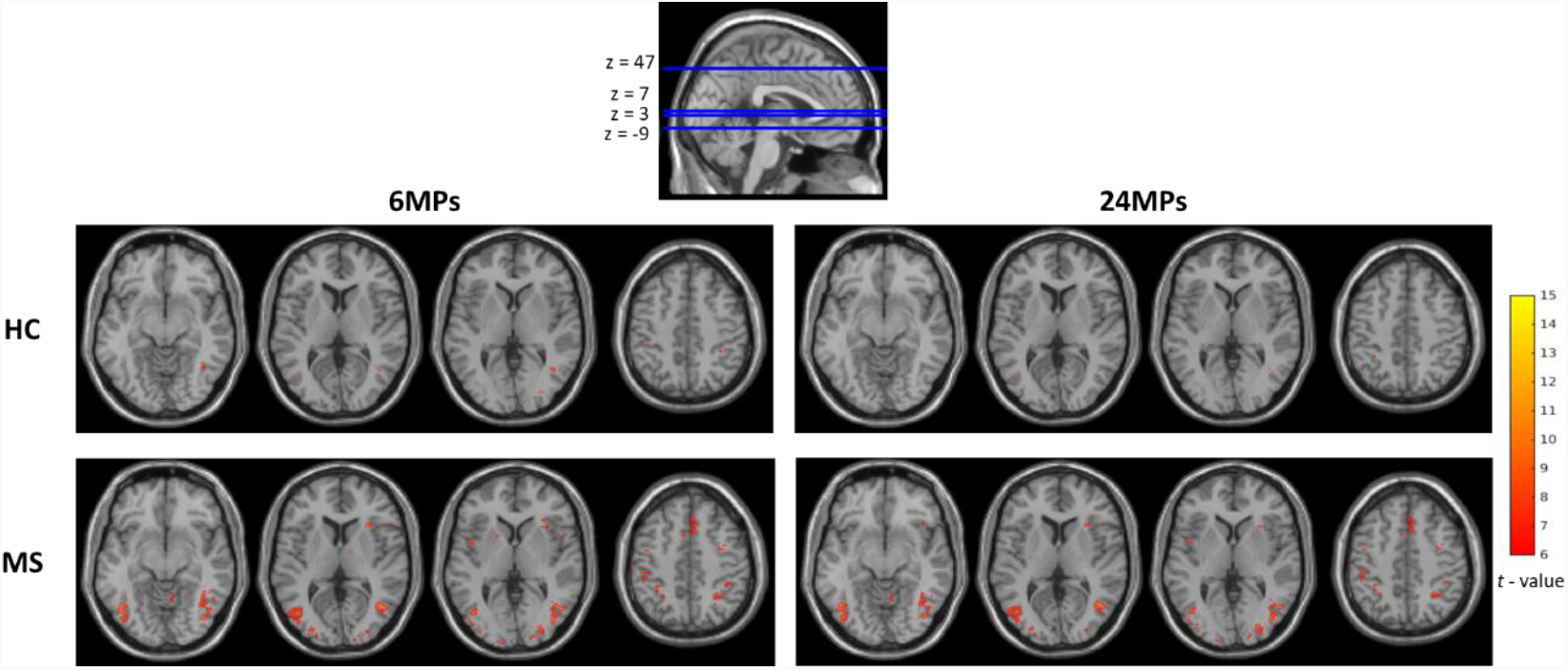
Group mean activation maps, resulting from the contrast [motion – no motion] in all runs. On the left are represented activation maps resulting from models with 6 MPs for each group. On the right are represented the activation maps resulting from models with 24 MPs for each group. Results are presented at a voxel *p*-value < 0.05, FWE corrected for multiple comparisons. Color bar scale represents t-values. The t-value is the result of the statistical test (t-test) in each voxel and measures the size of the difference calculated between the BOLD signal in the presence of motion stimuli and the BOLD signal during the absence of motion stimuli. The higher the t-value the most correlated is the BOLD signal with the specified contrast in a given brain region, thus more sensitive is that (group of) voxel(s). We can observe slightly higher extent of significant activations in the maps with 6 MPs.

The three-way interaction from the three-way mixed MANOVA was significant, *p* = 0.003, and the main effect of *Group* was also significant, *p* = 0.011 for Z-max and *p* = 0.018 for Z-mean, meaning that the group has influence on the correction approach. As the three-way interaction was significant, we fixed the groups and performed a two-way MANOVA in each (two within-subjects factors: *MPs* and *Correction Method*) to check what is the best approach for correction of gradual shifts and motion outliers in each group. In the MS group, the two-way interaction was non-significant, *p* = 0.607, which means that there aren’t significant differences between combinations of MPs and motion outliers’ correction method. Again, the main effect of *MPs* was significant, *p* < 0.001. For the HC group, the two-way interaction was significant, *p* = 0.002, which suggests that the combinations of the number of MPs with the motion outliers’ correction method might yield different results. Similarly, to the MS group, in the HC group the main effect of MPs was significant, p < 0.001. Following the rationale given by the first comparison, that using 6 MPs is better than using 24 MPs and given the fact that the main effect of MPs of these two MANOVAs performed in each group was significant, with pairwise comparisons showing higher Z-scores for 6 MPs in both groups, we proceeded with one-way MANOVAs (one within-subjects factor: *Correction Method*) in each group for models with 6 MPs to determine what is the best motion outliers’ correction method. In the MS group, the one-way MANOVA showed that the effect of *Correction Method* is significant, *p* = 0.013. Pairwise comparisons revealed that the best correction method seems to be volume interpolation, however differences between volume interpolation and FD are not statistically significant (*p* = 1). The difference between DVARS and FD is statistically significant, p = 0.011, with FD performing better than DVARS. In the HC group, the one-way MANOVA revealed that the effect of *Correction Method* was significant, *p* = 0.035. The pairwise comparisons showed that the best method was DVARS, however the difference between DVARS and volume interpolation is non-significant (*p* = 0.076). Also, the difference between DVARS and FD although significant is small (*p* = 0.046).

Figure 3 illustrates mean BOLD signal inside the Z-max cluster, located in the visual region hMT+, before and after motion correction and mean FD time courses for one participant of each group.

**Figure 3:**
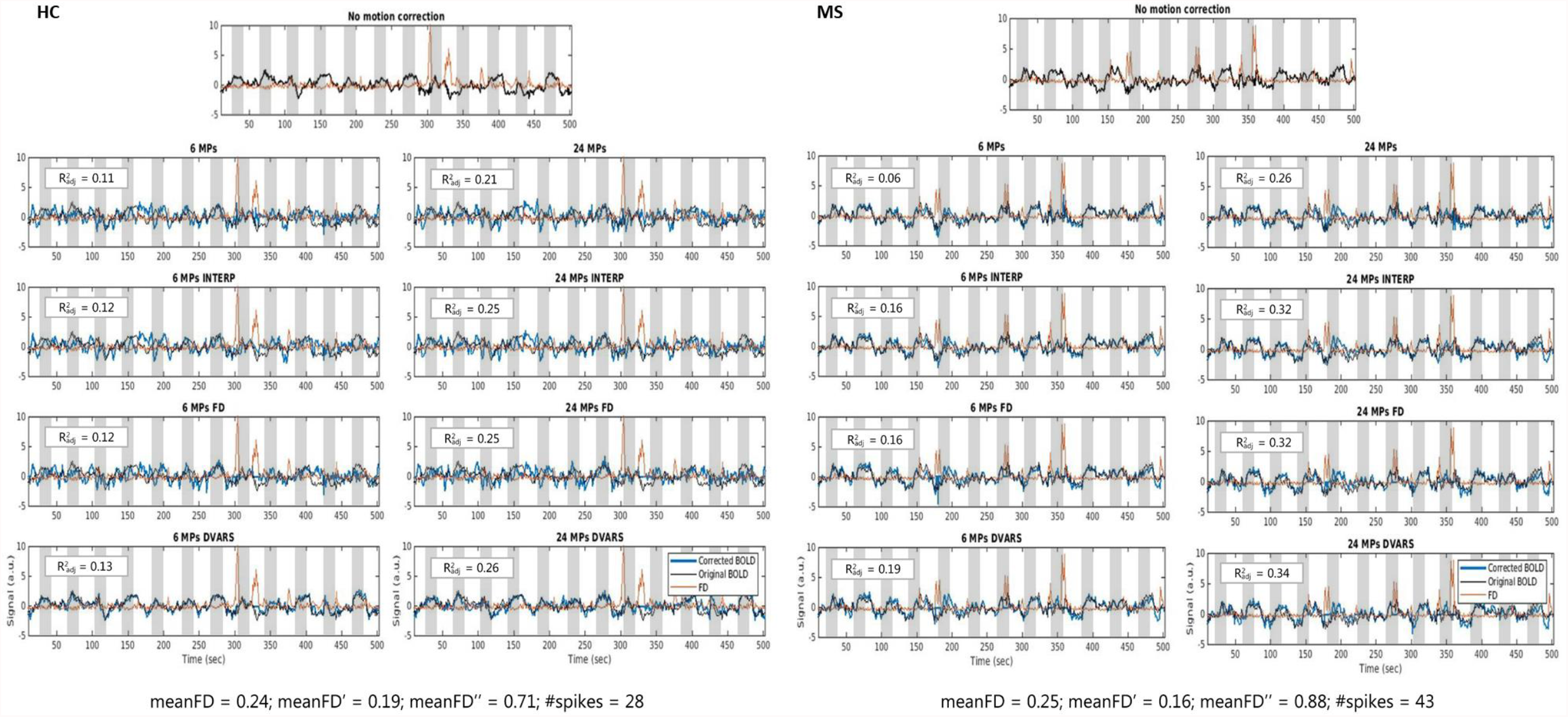
(left) Time courses of the BOLD signal, before motion correction and after motion correction with each approach, and mean FD for one participant from the HC group. (right) Time courses of the BOLD signal, before motion correction and after motion correction with each approach, and mean FD for one participant from the MS group. Measurements of FD allow the identification of spikes in the data, e.g., close to 300s (HC participant) and 350s (MS participant). Through the R^2^_adj_ formula, which indicates the amount of variance of the average BOLD signal without motion correction that is explained by motion regressors, we can compute how much motion effect is possible to remove with each correction method. The higher the values, the more motion contributions are removed, although too much variance might be removed with many motion parameters, including information not related to motion.

## 4. Discussion

There is a lack of consensus regarding which is the best approach to mitigate the effects of head motion in fMRI data. Reaching a consensus on the best strategy is even more important in the clinical context to produce reliable interpretations and foster clinical application. In this study we compared different strategies to compensate for head motion in fMRI data in a group of MS patients and a group of HC performing two visual tasks. We found that the group has influence on the choice of the motion correction approach, with 6 MPs combined with either volume interpolation or scrubbing with FD being the best correction approaches in the MS group, while 6 MPs combined with either scrubbing with DVARS or volume interpolation are the best correction approaches in the HC group.

We started by characterizing head motion in the two groups to study if the presence of disease affects motion occurrence. This comparison revealed that no significant differences in head motion were found between MS patients and HC, i.e., early diagnosed patients do not move more than the HC participants. While previous studies found clear evidence of greater motion in patients with MS than in HC subjects (Saccà et al., 2018; Wylie et al., 2014), our results may be due to the fact that the participants in this study are in early stages of the disease, have lower levels of EDSS and therefore do not show physical disabilities. Furthermore, in this study patients with MS are cognitively preserved, while others have investigated patients with cognitive impairment and suggest that healthy individuals and cognitively preserved patients with MS may process the cognitive task with enough efficiency that cerebral resources remain available for remaining still (Wylie et al., 2014). These authors have also shown a linear increase in movement of patients with MS and HC (to a less extent) as task difficulty increased. In this case, it might happen that our task is not demanding enough for these effects to stand out. Regarding the metrics we used to compare motion between groups, in addition to the well-known FD, we computed two variations of FD to investigate if the overall motion observed in each participant was mainly due to gradual head shifts or due the observation of motion outliers (abrupt motion spikes). The violin plots of these metrics show that values of FD without considering the points where motion outliers were detected are very similar to the conventional FD (all time points) values, supporting that in this cohort the abrupt movements of the head were not very problematic in overwhelmingly drive overall motion quantification. Nevertheless, it can be interesting and useful to check these motion metrics before deciding which correction approach should be put into practice.

Next, we compared the most commonly used strategies to correct the effects of head motion. In this comparison we aimed to study: 1) if the group has influence on the choice of the correction method; 2) if including temporal derivatives of MPs would improve the correction of gradual movements; 3) if models with only these MPs regressors would be enough to compensate for all the head motion, even for the more abrupt movements; 4) if not, which combination, between number of MPs and scrubbing with FD, scrubbing with DVARS and volume interpolation would correct better the effects of motion outliers. These analyses are particularly relevant in task-related and resting-state (rs) functional connectivity fMRI studies, which are rapidly increasing in clinical research (Goto et al., 2016). Particularly, MS is a disconnection disease that is due to structural damage but also functional connectivity alterations, with fMRI representing a gold-standard technique to investigate it (Tahedl et al., 2018). Previous studies have shown that group differences in head motion between control and patient groups cause group differences in the resting-state network with rs-fMRI (Lee et al., 2013; Maknojia et al., 2019; Saccà et al., 2019; Song et al., 2012). This raises the importance of these processing steps in functional connectivity studies, where one wants to study functionally connected networks throughout the brain that are correlated only due to the stimulation or cognitive processing, in task-based fMRI, or due their intrinsic functional organization at rest, not because of head motion. To our knowledge, there is a lack of this kind of studies in the MS context. No study has so far focused on determining the extent at which the effects of different motion correction approaches differ between groups in clinical studies of MS. Thus, more than reporting head motion differences, the investigation of which correction method is more suitable for these special cases is warranted, to obtain the most reliable results as possible.

The comparison between motion correction approaches considering only 6 or 24MPs, which are mainly designed to mitigate the effects of gradual head movements, revealed that higher Z-score values are obtained when considering 6 MPs regardless the group, which means that using 6 MPs is better than using 24 MPs. This suggests that task-specific brain regions are detected with higher sensitivity and less biased by noise due to a better motion correction when using 6 MPs relatively to using 24 MPs. The activation maps resultant from the GLM analysis show that maps with 6 MPs have larger activations (more activated voxels) than maps with 24 MPs. These results are consistent with literature reporting that adding temporal derivatives can result in loss of degrees of freedom and therefore loss of valuable information (Yang et al., 2019). In the context of this visual motion task, correcting head shifts with 6 MPs seems to be enough to cover the effects of gradual movements.

The traditional and common analyses use only a set of MPs as regressors to correct motion effects. However, to answer the question if models with only MPs regressors would be enough to compensate for all the head motion, even for the more abrupt movements, a third analysis was performed in which models with combinations of MPs and motion outliers’ correction methods were considered. The Z-score values are very similar between the models with only the MPs regressors and the rest of the models combining MPs with detection and correction of motion outliers. At a first glance this might mean that it’s not worth adding additional methods to compensate for the effects of more abrupt movements. However, actually because of that similarity and the fact that there is no considerable decrease in Z-values, which would suggest loss of valuable information, we consider that adding methods such as scrubbing, or volume interpolation are indeed crucial to eliminate residual noise and to compensate for putative motion outliers’ effects. The *R*^2^ values indicated in Figure 3 with an example of original and corrected time courses for one participant of each group also suggest that adding such methods help, indeed, to compensate for extra motion contributions without loss of signal of interest. This third analysis revealed that the motion correction approach differs between groups. This could be explained by the fact that motion outliers can happen at different moments in time between groups, e.g., if in one group the motion outliers always happen in periods of a stimulation condition, and in the other group always in fixation, this could lead to different corrections. Nonetheless, this is not the case in our study, as the number of motion outliers during stimulation or fixation periods between groups is approximately the same (described in supplementary material). More importantly, although the movement can be the same (in general, or even in outliers), the neuronal response behind the BOLD signal and its dynamics might be different between groups, so it wouldn’t be completely surprising that if the signal is by nature different between groups, then the motion correction of that signal might also yield different results. In fact, differences in neuronal responses behind the BOLD signal between MS and HC were recently described in the literature (Hubbard et al., 2016; Stickland et al., 2019; West et al., 2021). Activation maps with higher Z-scores ended up being those that result from models with 6 MPs and volume interpolation or 6 MPs and scrubbing with FD for the MS group, and models with 6 MPs and scrubbing with DVARS or 6 MPs and volume interpolation for the HC group. To our knowledge there are no studies of which strategy is best to correct motion outliers, with a direct comparison in the same data, between scrubbing, which is a modelling strategy, and volume interpolation, although the two approaches are widely used. Thus, it is important to discuss the impact of modelling motion outliers and interpolation in the data. Modelling motion outliers through scrubbing is a widely used technique to correct sudden movements of the head, however it creates temporal discontinuities. Interpolation overcomes this problem and avoids side effects in the high pass filtering step (Michielsen et al., 2011). However, volume interpolation induces synthetic data, and the duration of the censored segment, as well as the type of interpolation (linear, Fourier, wavelets or splines), may produce different effects that further depend on the choice of these parameters (Caballero-Gaudes and Reynolds, 2017). These effects and the negative impacts of using interpolation must be further investigated. This analysis also allowed us to directly compare the performance between motion outliers’ detection metrics, FD and DVARS. FD seems to perform better in the MS group while DVARS is preferable in the HC group. It is presently unclear whether one index captures data quality better than the other (Power et al., 2012). Indeed, the choice of one of them, given the ease of producing either measure, is left unresolved. Hence, there was a need to systematically compare them in the same data in combination with different number of MP regressors.

Apart from more traditional ways to deal with head motion, there are other techniques that can be implemented. Data driven strategies, namely algorithms such as PCA or ICA, which first decompose the data into a set of components, then the corrected fMRI data are obtained by removing the contribution of motion-related components (Caballero-Gaudes and Reynolds, 2017; Liu, 2016). Also, external optical tracking systems that constantly measure the position of the head or the use of dedicated sequences with navigator echoes or active markers are such examples (Caballero-Gaudes and Reynolds, 2017; Maknojia et al., 2019). However, here, we focused our study on these commonly and easily applicable methods, since we wanted to apply them in a clinical context where method implementation should be as easy and less time consuming as possible. We acknowledge the relatively limited sample size; thus, these results should be seen as suggestive regarding the recommendations to future studies. As to our knowledge, there are no fMRI studies with focus on MS to identify the best approach for correcting head motion, so this work is a first and crucial step towards this goal. Here, volume interpolation shows its relevance as it matches the performance of the common scrubbing methods, and results in the MS group suggest that volume interpolation may even outperform these methods. As MS can cause alterations in brain activity, which could influence the results, validation of these results is needed in future studies, namely in other healthy/patient cohorts alone with more data and considering other task designs. Nevertheless, with our results, we suggest that the optimal method, which reflects the best compromise between homogeneity of methodology between groups and performance, is the combination of 6 MPs with outliers’ interpolation. Moreover, this study represents a first step towards a more standard procedure for head motion correction in fMRI studies in this context.

## 5. Conclusions

In this study we characterized head motion in patients with early MS and healthy controls and compared different techniques to tackle head motion in task-based fMRI data to reach a consensus on the best strategies to use. While there were no differences between groups in motion quantification metrics, data analysis of quality metrics such as Z-score values were different between groups. Models with 6 MPs and volume interpolation or 6 MPs and scrubbing with FD were the best correction methods for the MS group, while models with 6 MPs and scrubbing with DVARS or 6 MPs and volume interpolation were the best correction methods for the HC group. This study is the first to systematically investigate the best approach for correcting head motion in MS, through comparison of commonly used and easy to implement approaches to correct head motion effects such as motion regression, scrubbing and volume interpolation. Our results pave the way towards finding an optimal motion correction strategy, which is required to improve the accuracy of fMRI analyses, crucially in clinical studies with patient populations.

## Data Availability

All data produced in the present study are available upon reasonable request to the authors.

## Acknowledgments

We would like to thank the participants for their involvement in this study. We are also very grateful to Sónia Afonso and Tânia Lopes for the help with MRI setup and scanning.

## Ethics approval

The study was approved by the Ethics Committee of the Faculty of Medicine of the University of Coimbra and the Ethics Committee of the Coimbra Hospital University Center and was conducted in accordance with the 1964 Declaration of Helsinki and its later amendments. All participants provided written informed consent to participate in the study.

## Material and Code availability

The datasets generated during and/or analyzed during the current study are available from the corresponding author on reasonable request.

## Supplementary Material

**Figure S1:**
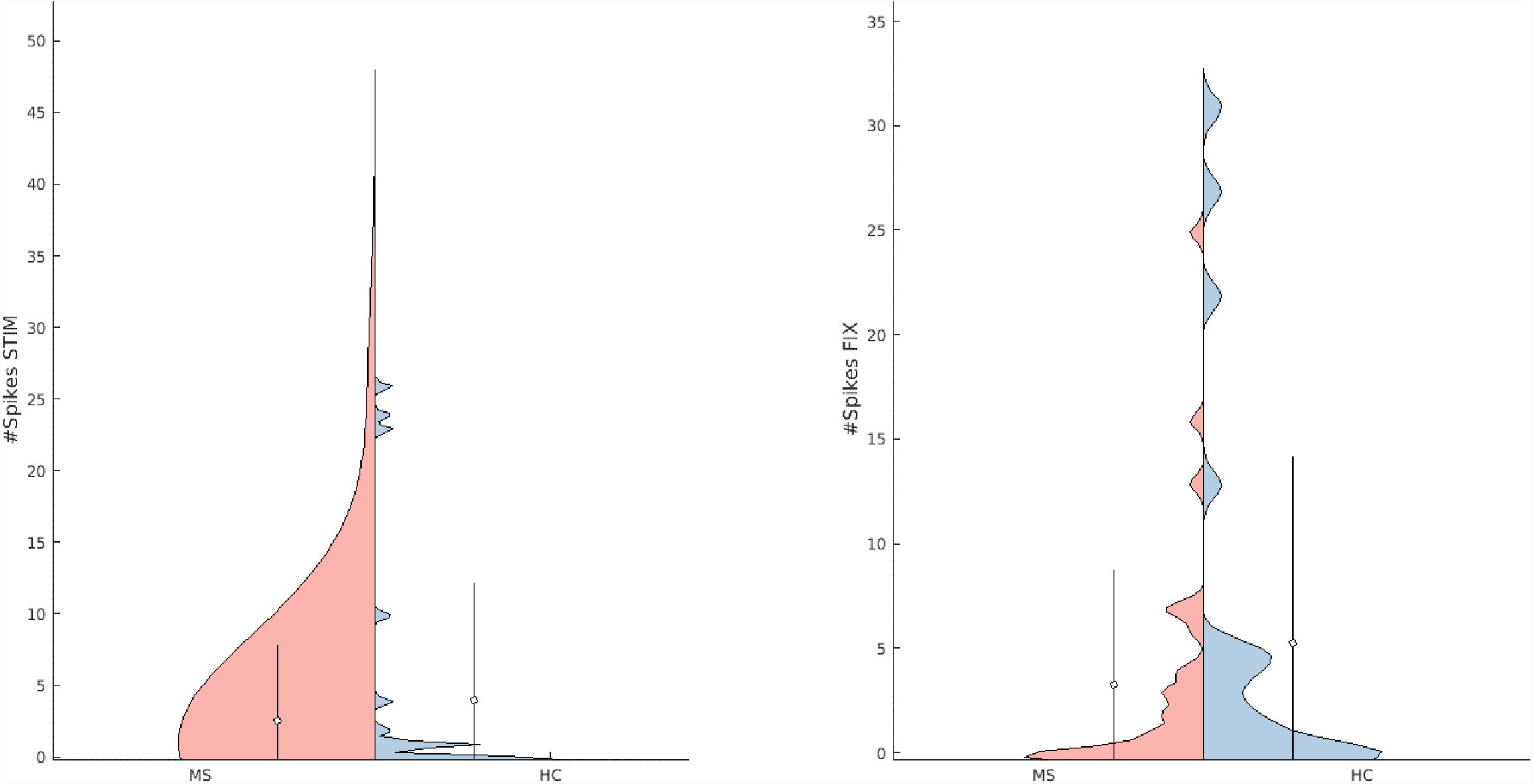
(left) Violin plots of number of motion outliers during stimulation periods. (right) Violin plots of number of motion outliers during fixation periods. Red and blue represent the groups of MS patients and HC, respectively. The dots and vertical lines in each group represent the mean ± standard deviation. Both distributions are quite similar, evidencing no differences in number of motion outliers during stimulation or fixation periods between groups, as supported by t-tests (*p*-value (#Spikes_STIM) = 0.84; *p*-value (#Spikes_FIX) = 0.60, corrected for multiple comparisons).

## References

Andersson, J.L., Skare, S., Ashburner, J., 2003. How to correct susceptibility distortions in spinecho echo-planar images: Application to diffusion tensor imaging. Neuroimage 20, 870– 888. https://doi.org/10.1016/S1053-8119(03)00336-7

Binnewijzend, M.A., Schoonheim, M.M., Sanz-Arigita, E., Wink, A.M., van der Flier, W.M., Tolboom, N., Adriaanse, S.M., Damoiseaux, J.S., Scheltens, P., van Berckel, B.N., Barkhof, F., 2012. Resting-state fMRI changes in Alzheimer’s disease and mild cognitive impairment. Neurobiol. Aging 33, 2018–2028. https://doi.org/10.1016/j.neurobiolaging.2011.07.003

Boonstra, F., Florescu, G., Evans, A., Steward, C., Mitchell, P., Desmond, P., Moffat, B., Butzkueven, H., Kolbe, S., van der Walt, A., 2017. Tremor in multiple sclerosis is associated with cerebello-thalamic pathology. J. Neural Transm. 124, 1509–1514. https://doi.org/10.1007/s00702-017-1798-4

Caballero-Gaudes, C., Reynolds, R.C., 2017. Methods for cleaning the BOLD fMRI signal. Neuroimage 154, 128–149. https://doi.org/10.1016/j.neuroimage.2016.12.018

Ciric, R., Wolf, D.H., Power, J.D., Roalf, D.R., Baum, G., Ruparel, K., Shinohara, R.T., Elliott, M.A., Eickhofff, S.B., Davatzikos, C., Gur, R.C., Gur, R.E., Bassett, D.S., Satterthwaite, T.D., 2017. Benchmarking of participant-level confound regression strategies for the control of motion artifact in studies of functional connectivity. Neuroimage 154, 174–187. https://doi.org/10.1016/j.neuroimage.2017.03.020

Di, X., Gohel, S., Kim, E.H., Biswal, B.B., 2013. Task vs. rest-different network configurations between the coactivation and the resting-state brain networks. Front. Hum. Neurosci. 7, Article 493. https://doi.org/10.3389/fnhum.2013.00493

Eijlers, A.J., Wink, A.M., Meijer, K.A., Douw, L., Geurts, J.J., Schoonheim, M.M., 2019. Reduced network dynamics on functional MRI signals cognitive impairment in multiple sclerosis. Radiology 292, 449–457. https://doi.org/10.1148/radiol.2019182623

Goto, M., Abe, O., Miyati, T., Yamasue, H., Gomi, T., Takeda, T., 2016. Head motion and correction methods in resting-state functional MRI. Magn. Reson. Med. Sci. 15, 178–186. https://doi.org/10.2463/mrms.rev.2015-0060

Griffanti, L., Rolinski, M., Szewczyk-Krolikowski, K., Menke, R.A., Filippini, N., Zamboni, G., Jenkinson, M., Hu, M.T.M., Mackay, C.E., 2016. Challenges in the reproducibility of clinical studies with resting state fMRI: An example in early Parkinson’s disease. Neuroimage 124, 704–713. https://doi.org/10.1016/j.neuroimage.2015.09.021

Hubbard, N.A., Turner, M., Hutchison, J.L., Ouyang, A., Strain, J., Oasay, L., Sundaram, S., Davis, S., Remington, G., Brigante, R., Huang, H., Hart, J., Frohman, T., Frohman, E., Biswal, B.B., Rypma, B., 2016. Multiple sclerosis-related white matter microstructural change alters the BOLD hemodynamic response. J. Cereb. Blood Flow Metab. 36, 1872– 1884. https://doi.org/10.1177/0271678X15615133

Kasper, L., Bollmann, S., Diaconescu, A.O., Hutton, C., Heinzle, J., Iglesias, S., Hauser, T.U., Sebold, M., Manjaly, Z.M., Pruessmann, K.P., Stephan, K.E., 2017. The PhysIO Toolbox for Modeling Physiological Noise in fMRI Data. J. Neurosci. Methods 276, 56–72. https://doi.org/10.1016/j.jneumeth.2016.10.019

Langdon, D.W., Amato, M.P., Boringa, J., Brochet, B., Foley, F., Fredrikson, S., Hämäläinen, P., Hartung, H.-P., Krupp, L., Penner, I.K., Reder, A.T., Benedict, R.H., 2012. Recommendations for a brief international cognitive assessment for multiple sclerosis (BICAMS). Mult. Scler. J. 18, 891–898. https://doi.org/10.1177/1352458511431076

Lee, M.H., Smyser, C.D., Shimony, J.S., 2013. Resting state fMRI: A review of methods and clinical applications. AJNR Am J Neuroradiol 34, 1866–1872. https://doi.org/10.3174/ajnr.A3263

Liu, T.T., 2016. NeuroImage Noise contributions to the fMRI signal : An overview. Neuroimage 143, 141–151. https://doi.org/10.1016/j.neuroimage.2016.09.008

Lowe, M.J., Horenstein, C.I., Bedekar, D., Marie, R.A., Stone, L., Bhattacharyya, P.K., Dzemidzic, M., Phillips, M.D., 2006. Cortical Activation Volume During a Bilateral Motor Task in Multiple Sclerosis Patient : A Study of the Effect of Subject. Dimens. Contemp. Ger. Arts Lett. D, 2314–2314.

Maknojia, S., Churchill, N.W., Schweizer, T.A., Graham, S., 2019. Resting state fMRI: Going through the motions. Front. Neurosci. 13, 1–13. https://doi.org/10.3389/fnins.2019.00825

Mascali, D., Moraschi, M., DiNuzzo, M., Tommasin, S., Fratini, M., Gili, T., Wise, R.G., Mangia, S., Macaluso, E., Giove, F., 2021. Evaluation of denoising strategies for task-based functional connectivity: Equalizing residual motion artifacts between rest and cognitively demanding tasks. Hum. Brain Mapp. 1805–1828. https://doi.org/10.1002/hbm.25332

Mazaika, P.K., Hoeft, F., Glover, G.H., Reiss, A.L., 2009. Methods and Software for fMRI Analysis for Clinical Subjects.

Mckechanie, A.G., Campbell, S., Eley, S.E.A., Stanfield, A.C., 2019. Autism in Fragile X Syndrome ; A Functional MRI Study of Facial Emotion-Processing. Genes (Basel).

Meijer, K.A., Steenwijk, M.D., Douw, L., Schoonheim, M.M., Geurts, J.J., 2020. Long-range connections are more severely damaged and relevant for cognition in multiple sclerosis. Brain 143, 150–160. https://doi.org/10.1093/brain/awz355

Michielsen, M.E., Selles, R.W., Geest, J.N. Van Der, Eckhardt, M., Yavuzer, G., Stam, H.J., Smits, M., Ribbers, G.M., Bussmann, J.B.J., 2011. Motor Recovery and Cortical Reorganization After Mirror Therapy in Chronic Stroke Patients : A Phase II Randomized Controlled Trial. Neurorehabil. Neural Repair 25, 223–233. https://doi.org/10.1177/1545968310385127

Montgomery, D.C., Peck, E.A., Vining, G.G., 2012. Introduction to Linear Regression Analysis. John Wiley & Sons, Ltd.

Parkes, L., Fulcher, B., Yücel, M., Fornito, A., 2018. An evaluation of the efficacy, reliability, and sensitivity of motion correction strategies for resting-state functional MRI. Neuroimage 171, 415–436. https://doi.org/10.1016/j.neuroimage.2017.12.073

Pernet, C.R., 2014. Misconceptions in the use of the General Linear Model applied to functional MRI: a tutorial for junior neuro-imagers. Front. Neurosci. 8, Article 1. https://doi.org/10.3389/fnins.2014.00001

Power, J.D., Barnes, K. a., Snyder, A.Z., Schlaggar, B.L., Petersen, S.E., 2012. Spurious but systematic correlations in functional connectivity MRI networks arise from subject motion. Neuroimage 59, 2142–2154. https://doi.org/10.1016/j.neuroimage.2011.10.018

Power, J.D., Mitra, A., Laumann, T.O., Snyder, A.Z., Schlaggar, B.L., Petersen, S.E., 2014. Methods to detect, characterize, and remove motion artifact in resting state fMRI. Neuroimage 84. https://doi.org/10.1016/j.neuroimage.2013.08.048

Power, J.D., Schlaggar, B.L., Petersen, S.E., 2015. Recent progress and outstanding issues in motion correction in resting state fMRI. Neuroimage 105, 536–551. https://doi.org/10.1016/j.neuroimage.2014.10.044

Roosendaal, S.D., Schoonheim, M.M., Hulst, H.E., Sanz-Arigita, E.J., Smith, S.M., Geurts, J.J.G., Barkhof, F., 2010. Resting state networks change in clinically isolated syndrome. Brain 133, 1612–1621. https://doi.org/10.1093/brain/awq058

Rudas, J., Martı, D., Castellanos, G., Demertzi, A., Martial, C., Carrie, M., 2020. Time-Delay Latency of Resting-State Blood Oxygen Level-Dependent Signal Related to the Level of Consciousness in Patients with Severe Consciousness Impairment. Brain Connect. 10, 83– 94. https://doi.org/10.1089/brain.2019.0716

Saccà, V., Sarica, A., Novellino, F., Barone, S., Filippelli, E., Granata, A., Demonte, G., Nisticò, R., Valentino, P., Quattrone, A., 2018. Studying of Resting State fMRI head movements in Multiple Sclerosis and Essential Tremor patients 69, 2018.

Saccà, V., Sarica, A., Novellino, F., Barone, S., Valentino, P., Quattrone, A., 2019. Evaluation of the Subject Involuntary Head Motions in rs-fMRI Acquisition : Characterization of EarlyMS Movements, in: Società Italiana Di Neonatologia - 50° Congresso Nazionale.

Saccà, V., Sarica, A., Quattrone, Andrea, Rocca, F., Quattrone, Aldo, Novellino, F., 2021. Aging effect on head motion: A Machine Learning study on resting state fMRI data. J. Neurosci. Methods 352. https://doi.org/10.1016/j.jneumeth.2021.109084

Satterthwaite, T.D., Elliott, M.A., Gerraty, R.T., Ruparel, K., Loughead, J., Calkins, M.E., Eickhoff, S.B., Hakonarson, H., Gur, R.E.R.C., Gur, R.E.R.C., Wolf, D.H., 2013. An improved framework for confound regression and filtering for control of motion artifact in the preprocessing of resting-state functional connectivity data. Neuroimage 64, 240–256. https://doi.org/10.1016/j.neuroimage.2012.08.052

Sbardella, E., Petsas, N., Tona, F., Pantano, P., 2015. Resting-state fMRI in MS: General concepts and brief overview of its application. Biomed Res. Int. Article ID 212693. https://doi.org/10.1155/2015/212693

Schoonheim, M.M., Geurts, J.J., Barkhof, F., 2010. The limits of functional reorganization in multiple sclerosis. Neurology 74, 1246–1247. https://doi.org/10.1212/WNL.0b013e3181db9957

Schoonheim, M.M., Meijer, K.A., Geurts, J.J., 2015. Network collapse and cognitive impairment in multiple sclerosis. Front. Neurol. 6, 1–5. https://doi.org/10.3389/fneur.2015.00082

Seto, E., Sela, G., McIlroy, W., Black, E., Staines, W., Bronskill, M., McIntosh, A., Graham, S., 2001. Quantifying head motion associated with motor tasks used in fMRI. Neuroimage 14, 284–297. https://doi.org/10.1006/nimg.2001.0829

Shu, N., Duan, Y., Xia, M., Schoonheim, M.M., Huang, J., Ren, Z., Sun, Z., Ye, J., Dong, H., Shi, F.D., Barkhof, F., Li, K., Liu, Y., 2016. Disrupted topological organization of structural and functional brain connectomes in clinically isolated syndrome and multiple sclerosis. Sci. Rep. 6, 1–11. https://doi.org/10.1038/srep29383

Siegel, J.S., Power, J.D., Dubis, J.W., Vogel, A.C., Church, J.A., Schlaggar, B.L., Petersen, S.E., 2014. Statistical Improvements in Functional Magnetic Resonance Imaging Analyses Produced by Censoring High-Motion Data Points. Hum. Brain Mapp. 1981–1996. https://doi.org/10.1002/hbm.22307

Song, J., Desphande, A.S., Meier, T.B., Tudorascu, D.L., Vergun, S., Nair, V.A., Biswal, B.B., Meyerand, M.E., Birn, R.M., Bellec, P., Prabhakaran, V., 2012. Age-Related Differences in Test-Retest Reliability in Resting-State Brain Functional Connectivity. PLoS One 7, 1–16. https://doi.org/10.1371/journal.pone.0049847

Stickland, R., Allen, M., Magazzini, L., Singh, K.D., Wise, R.G., Tomassini, V., 2019. Neurovascular Coupling During Visual Stimulation in Multiple Sclerosis: A MEG-fMRI Study. Neuroscience 403, 54–69. https://doi.org/10.1016/j.neuroscience.2018.03.018

Tahedl, M., Levine, S.M., Greenlee, M.W., Weissert, R., Schwarzbach, J. V., 2018. Functional connectivity in multiple sclerosis: Recent findings and future directions. Front. Neurol. 9, 1–18. https://doi.org/10.3389/fneur.2018.00828

Thompson, A.J., Banwell, B.L., Barkhof, F., Carroll, W.M., Coetzee, T., Comi, G., Correale, J., Fazekas, F., Filippi, M., Freedman, M.S., Fujihara, K., Galetta, S.L., Hartung, H.P., Kappos, L., Lublin, F.D., Marrie, R.A., Miller, A.E., Miller, D.H., Montalban, X., Mowry, E.M., Sorensen, P.S., Tintoré, M., Traboulsee, A.L., Trojano, M., Uitdehaag, B.M., Vukusic, S., Waubant, E., Weinshenker, B.G., Reingold, S.C., Cohen, J.A., 2018. Diagnosis of multiple sclerosis: 2017 revisions of the McDonald criteria. Lancet Neurol. 17, 162–173. https://doi.org/10.1016/S1474-4422(17)30470-2

Turner, R., Friston, K.J., Williams, S., Howard, R., Frackowiak, R.S.J., 1996. Movement-Related Effects in fMRI Time-Series. Magn. Reson. Med. 35, 346–355.

van Duinkerken, E., Schoonheim, M.M., Sanz-Arigita, E.J., IJzerman, R.G., Moll, A.C., Snoek, F.J., Ryan, C.M., Klein, M., Diamant, M., Barkhof, F., 2012. Resting-state brain networks in type 1 diabetic patients with and without microangiopathy and their relation to cognitive functions and disease variables. Diabetes 61, 1814–1821. https://doi.org/10.2337/db11-1358

West, K.L., Sivakolundu, D.K., Zuppichini, M.D., Turner, M.P., Spence, J.S., Lu, H., Okuda, D.T., Rypma, B., 2021. Altered task-induced cerebral blood flow and oxygen metabolism underlies motor impairment in multiple sclerosis. J. Cereb. Blood Flow Metab. 41, 182–193. https://doi.org/10.1177/0271678X20908356

Wylie, G.R., Genova, H., Deluca, J., Chiaravalloti, N., Sumowski, J.F., 2014. Functional magnetic resonance imaging movers and shakers: Does subject-movement cause sampling biasã Hum. Brain Mapp. 35, 1–13. https://doi.org/10.1002/hbm.22150

Yang, Z., Zhuang, X., Sreenivasan, K., Mishra, V., 2019. Robust Motion Regression of Resting-State Data Using a Convolutional Neural Network Model. Front. Neurosci. 13, 1–14. https://doi.org/10.3389/fnins.2019.00169

Zaitsev, M., Maclaren, J., Herbst, M., 2015. Motion artifacts in MRI: A complex problem with many partial solutions. J. Magn. Reson. Imaging 42, 887–901. https://doi.org/10.1002/jmri.24850

Zeng, L.L., Wang, D., Fox, M.D., Sabuncu, M., Hu, D., Ge, M., Buckner, R.L., Liu, H., 2014. Neurobiological basis of head motion in brain imaging. Proc. Natl. Acad. Sci. U. S. A. 111, 6058–6062. https://doi.org/10.1073/pnas.1317424111

